# The Repercussion of SARS-CoV-2 on the Blood Glucose Level of Diabetes Patients’ Prior and During the Lockdown in Bangladesh

**DOI:** 10.1101/2021.10.09.21264800

**Authors:** Shams Mohammad Abrar

## Abstract

**Purpose:** Diabetes Mellitus (DM) patients were exposed to subacute risk as a result of the unanticipated lockdown. Furthermore, most DM patients were unable to engage in physical activity during that period. This impediment to proper healthcare management had increased Blood Glucose Levels (BGL). Therefore, initiatives must be adopted to prevent the same result in the second lockdown in 2021 for the well-being of patients.

**Method:** This statistical analysis aimed to assess the rise in BGL of diabetic patients before and during the lockdown. A survey was conducted among the DM patients in the Bangladeshi cohort, who came from various socioeconomic backgrounds and included both men and women. The statistical modeling, performed with the help of stat-ease software, was conducted by applying the Analysis of Variance (ANOVA) method to Response Surface Methodology (RSM).

**Result:** Out of the 3 models applied (quadratic, main effect and sequential sum of squares for 2 factor interaction (2FI)) in 2 different response vectors the 2FI model was the best suited (p value - 0.0441 and 0.0015). The results yielded by the 2FI model were used to evaluate RSM.

**Conclusion:** The analysis had shown a significant rise in the BGL among the DM patients during the lockdown, and the patients with the higher BMI tend to have a more significant increment in the BGL. Male patients experienced a greater rise in BGL. Furthermore, elderly patients with high Random Blood Glucose (RBG) levels before lockdown were more likely to have high RBG levels during the lockdown.

## 1. Introduction

The pandemic of COVID-19 harmed the global health care system and the economy. Furthermore, it has created enormous turbulence in people’s daily lives. It has resulted in such a disaster that it is dubbed as Black Swan [1]. The SARS-CoV-2 outbreak was so severe that governments in various countries were forced to implement lockdown - enforcing border shutdowns, travel restrictions, and quarantine.

Diabetes, a devastating incurable health complication, is caused by the incapability of beta cells in the pancreas to produce adequate insulin for the body to utilize, leading to chronic high levels of blood glucose [2]. Globally, people aged between 20- and 79-years having diabetes are approximately 463 million, and impaired glucose tolerance is also 374 million [3]. In 2019 around 9.3% of the world’s population will be diagnosed with diabetes [4]. Also, the prevalence of DM among the middle class and lower-class families in developing countries is alarmingly high [5]. Similarly, Bangladesh is no stranger to the global epidemic of DM. The International Diabetes Federation (IDF) estimated that around 8.4 million people of Bangladesh out of 160 million have DM. Their study also implies that this number might double within 2025 [6]. DM is not only a long-time chronic disease, but also it brings forth many comorbidities among its patients. However, if the high BGL of the diabetes patients are controlled and disciplinary enactments of daily lifestyle are implemented, then most of the comorbidities of DM could be avoided [7].

Hyperglycemia, high BGL, is caused due to the failure of the pancreas to produce sufficient insulin [1]. Physical inactivity and obesity are one of the main reasons for hyperglycemia among DM patients. As a consequence, hyperglycemia may accompany growth impairment and proneness to certain infections, and acute hyperglycemia with ketoacidosis or the nonketotic hyperosmolar syndrome [8]. Meanwhile, due to the outbreak of SARS-CoV-2, the DM patients were unable to conduct their regular physical activity. Also, many DM patients were demotivated to continue their healthy diet.

The author was motivated by the substantial increment of BGL during lockdown among DM patients, and statistical analysis was done to analyze the effect on the blood sugar of DM patients from diverse socio-economic backgrounds. Considering the extensive plans taken by the Bangladesh government to obtain the status of a developed country in 20 years (2041) [9]. The software used for this approach was Stat-Ease by Design Expert. For statistical analysis, here, the implementation of the sequential sum of squares is conducted.

## 2. Methods

### 2.1 Data Collection

Recently, A survey was taken (voluntarily) on randomized 50 DM patients about their health conditions before and during the lockdown. As this is first-of-its-kind research as per the author’s knowledge and the pandemic was still ongoing during that period, the author could not collect more data from other outpatients.

### 2.2 Ethical Statement

The survey was conducted among the visiting/inpatients of Shaheed Khalek Ibrahim General Hospital right after the lockdown in 2021. The full acknowledgement goes to the patients who had voluntarily participated in the survey. The survey is ethically approved by the National Healthcare Network. This was solo research by the author himself. Only accurate results (for BP, FBG, RBG) are recorded in their checkups to doctors by the patients, from time to time every month, in their notepad.

### 2.3 Model Attributes

The feature vectors - the independent variables - for this statistical analysis were age, height, BMI, diabetes duration, Blood Pressure (BP), and blood sugar before lockdown. The total expenditure of elderly DM patients is higher than the young DM patients [10-12], and patients who have higher diabetic duration were also incurred to higher diabetes expenditure [13-16].

The Ideal BP for a healthy human being is 120/80 [17]. The ideal blood pressure for a diabetic individual is 135/85 mmHg, yet this value should not drop below 130/80 mmHg [18]. The patients were also asked about their BGL before the lockdown. The response was accounted for in two terms, the first was the Fasting Blood Glucose (FBG) and the second was the Random Blood Glucose (RBG).

### 2.4 Dependent Variable

The author recorded the patients’ FBG during the lockdown, i.e., pre-prandial blood glucose and RBG of each patient during the lockdown. The ideal FBG level should be <5.6 mmol/L, and the RBG level should be <7.8 mmol/L [19]. According to the medical news today, the FBG level of a diabetes patient should be ranging from 4.4 to 7.2. Subsequently, the RBG of diabetes patients should be less than ten mmol/L [20]. The values taken for features and response vectors were during the end of the lockdown, except for FBG and RBG before the lockdown.

### 2.5 Summary of the data

To get the overall summary and show the diversity of the data, Pandas, a python library for data analysis and manipulation, was used. From TABLE 1, it is seen that the mean age and BMI of the patients of the survey were approximately 47 years and 26.5, respectively. This indicates that most of the patients were overweight or not in the ideal BMI index. In Table-1, the FBG and RBG of patients during the lockdown - FBG (lock) and RBG (lock) respectively - was more than the RBG of patients before the lockdown. The table shows that the mean FBG and RBG of patients had increased during the lockdown.

**Table 1:** Summary of the Data using Pandas for different independent variables.

| Factor | Age | BMI | Diabetes Duration (DD) | SBP | DBP | FBG (lock) | RBG (lock) | FBG | RBG |
| --- | --- | --- | --- | --- | --- | --- | --- | --- | --- |
| **mean** | 47.43 | 26.54 | 11.4 | 131.00 | 92.45 | 12.13 | 16.91 | 8.33 | 11.82 |
| **\*std** | 10.6 | 3.68 | 9.67 | 20.72 | 11.52 | 3.29 | 5.27 | 2.33 | 3.94 |
| **min** | 29 | 20 | 0 | 90.00 | 70 | 6 | 7.80 | 5.10 | 7.10 |
| **25%** | 39.8 | 24.35 | 5 | 120 | 80 | 10 | 14 | 6.125000 | 9.25 |
| **50%** | 46 | 26.35 | 7 | 130 | 95 | 12.5 | 17.00 | 8.00 | 11.00 |
| **75%** | 56 | 27.60 | 15 | 140 | 100 | 14.75 | 20.00 | 9.75 | 13.75 |
| **max** | 72 | 39.2 | 40 | 200 | 110 | 21 | 25.00 | 15.00 | 27.00 |

\*std = Standard deviation
\*FBG (lock) = FBG during the lockdown
\*RBG (lock) = RBG during the lockdown
\*SBP = Systolic Blood Pressure
\*DBP = Diastolic Blood Pressure

### 2.6 Application of ANOVA

The empirical models were developed with the design procedures and Response Surface Methodology (RSM). The R^2^ value is the measure of the amount of variance with the mean and response-feature vector of the model, but it does not determine the optimum model for evaluation. In general, the higher the R-squared value indicates the better fit of the model. On the contrary, adjusted *R*^*2*^ is another modified version of R-squared in which it is adjusted around the degree of freedom. The reliability of the correlation is determined through the *R*^*2*^ adjusted value. Approximately, an *R*^*2*^ value greater than 0.75 is ideal for the model.

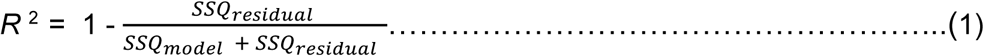

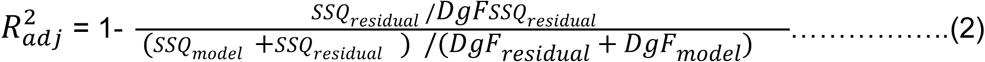

The responses of FBG and RBG during and after the lockdown were analyzed, and this was the indicator of the performance of the system. To analyze the model, the sequential sum of squares model (2FI) was used. For the design of the percentage of the FBG and RBG, the following mathematical relation was composed, i.e.

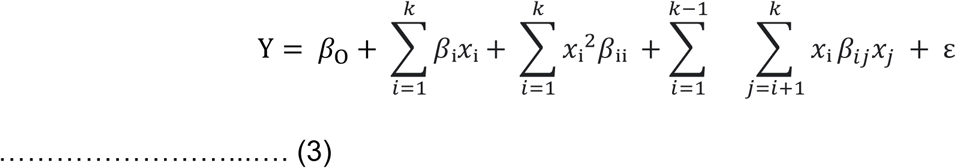

Here Y and ε is the response and error vector of the model respectively. Analysis of variance (ANOVA) and *F*-value analysis are used for the testing of the consequence of the second-order regression models. The *F*-value calculation can be expressed using the following equation.

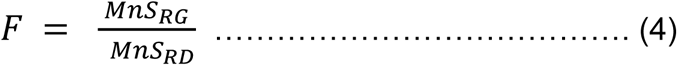

The DgF based *F* distribution for residuals is applied to calculate the *F*-value in the definite point of significance. *p-*value was used to analyze a regression coefficient. The extent of error is indicated by the coefficient of variation (*CV*) which is measured as the percentage of standard deviation over the mean value and given as

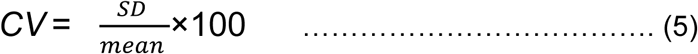

### 2.7 Model Fitting

This fit summary is necessary as it is used to determine the correct starting point for the final model. In Table 2, the summary of the 2FI, quadratic, main effect models for FBG and RBG are shown.

**Table 2:** Summary of sequential models for FBG and RBG during lockdown with statistical parameters.

| Source | Factor | Sum of squares | Degree of freedom | Mean square | F-value | p-value | R<sup>2</sup> |
| --- | --- | --- | --- | --- | --- | --- | --- |
| 2FI | FBG | 402.4 | 36 | 11.2 | 2.43 | 0.0441 (significant) | 0.87 |
|  | RBG | 1228.6 | 36 | 34.13 | 5.1 | 0.0015 (significant) | 0.94 |
| Main effect model | FBG | 194.5 | 8 | 24.3 | 3.73 | 0.0024 (significant) | 0.43 |
|  | RBG | 760.1 | 8 | 95.01 | 7.01 | <0.0001 (significant) | 0.58 |
| Quadratic | FBG | 248.2 | 43 | 5.77 | 1.73 | 0.253 | 0.56 |
|  | RBG | 733.45 | 43 | 17.06 | 3.47 | 0.0605 | 0.37 |

The small *p-value* (Prob > *F*), is the result when considering the null hypothesis as true, and a very small *p-value* indicates that the null hypothesis could be avoided. The *F*-value is a test for comparing data from the dataset by its mean square to its residual mean square. From Table 2, it is seen that the 2FI is a statistically significant model that is the most suited for FBG and RBG analysis; though it does not have the lowest *p*-value, its overall result is the most accurate. This is due to its *R*^*2*^ value being significantly higher than any other model while remaining significant. Secondly, the main effect model was the 2nd most significant - as it had the least *p*-value. The quadratic model was the least significant among the 3 models. As a result, the 2FI model is used for further analysis such as model graphs, ANOVA and diagnostics.

### 2.8 Fit statistics

The design expert software also generated coded equations (set by default to its high levels of the factor as +1 and low levels as -1) for both FBG and RBG during the lockdown in linear regression form. This predicted equation was used to evaluate the predicted *R*^*2*^ and the Adjusted *R*^*2*^ values.

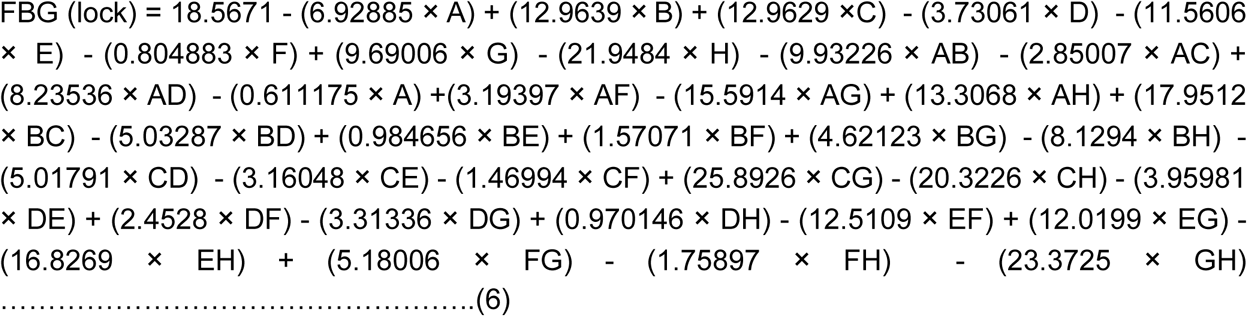

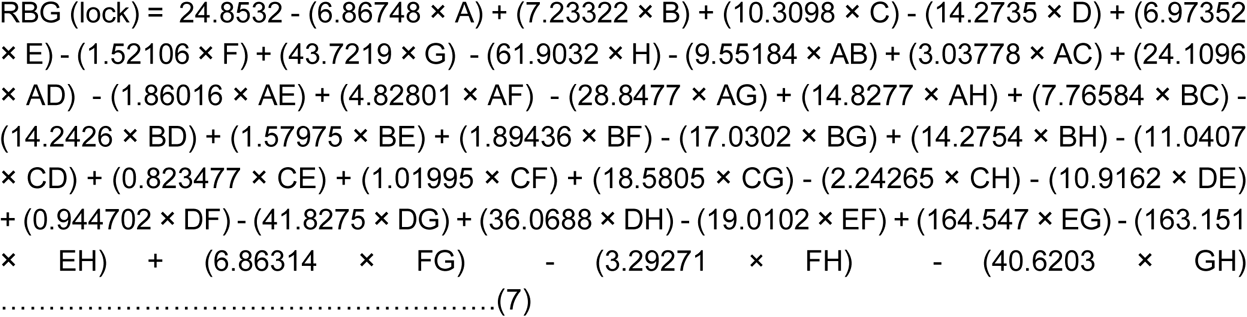

In the fit statistics of Table 3 and 4, the *% CV* is observed 17.22% for FBG analysis and 14.90% for RBG analysis. *CV*<10 is very good, 10-20 is good, 20-30 is acceptable, and *CV*>30 is not acceptable. Therefore, the *CV*% for the 2FI model is good enough for further analysis. It is seen that the models for FBG and RBG analysis have an *R*^*2*^*-*value of greater than 0.85 (0.87 for FBG (lock) and 0.94 for RBG (lock)). Likewise, the regression analysis was accurate for the model.

**Table 3:**
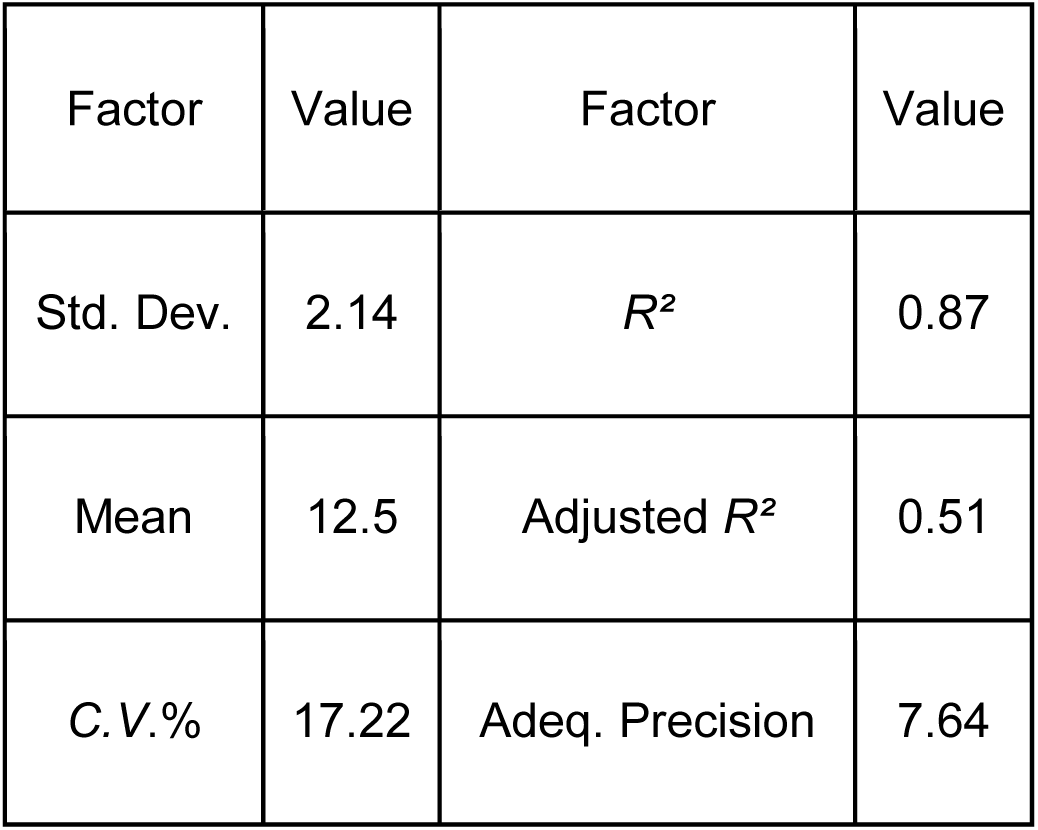
Fit statistics of FBG analysis during the lockdown.

**Table 4:**
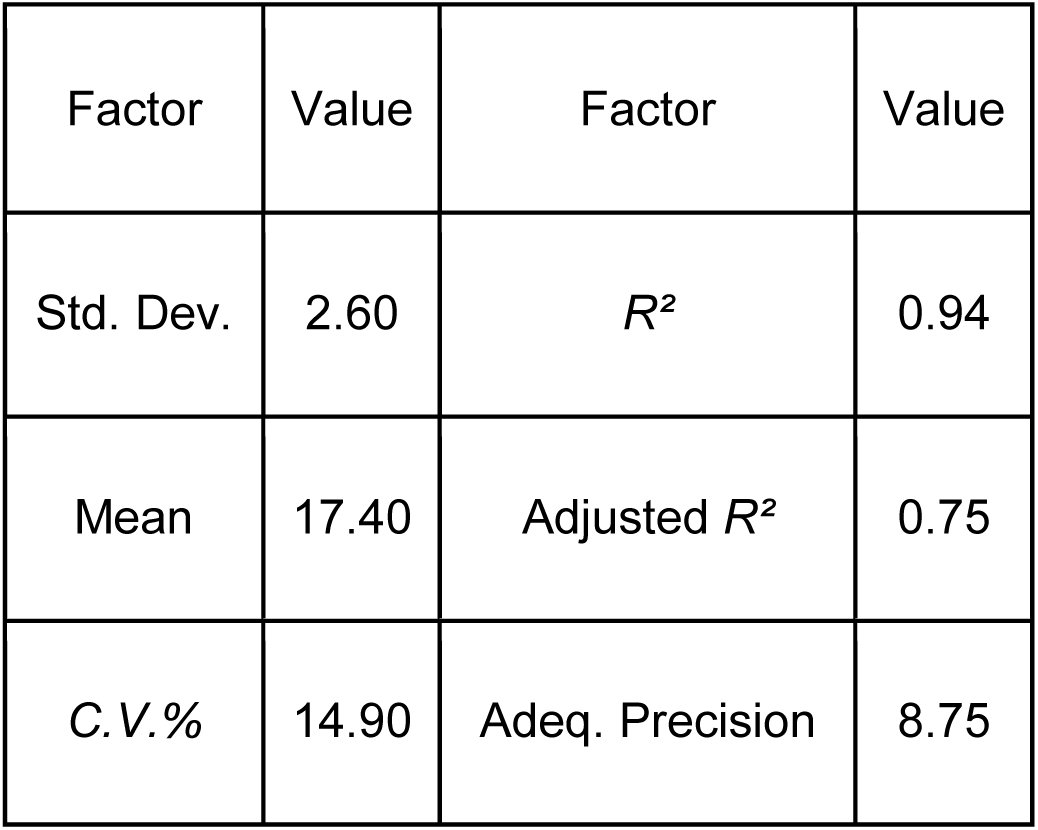
Fit statistics for RBG analysis during the lockdown.

### 2.9 Model statistics by diagnostic statistics

In this research, for checking the adequacy some diagnostic tools were adopted to check the results and process parameters. Models’ appropriateness was estimated implementing residuals (the difference between the actual value and residual value) and influenced plots - which determined empirical data coefficient in this research work. Components of variations are considered as residuals, usually, in the model, they are fitted imprecisely. It is predicted that its behavior is like a normal distribution feature - it is used to indicate whether the residuals follow a linear relationship with the distribution. For considering this research, as a proper method, a normal probability plot and graphical visualization were also taken as a methodology. The normal probability plot for externally studentized residuals and normal %values for FBG and RBG during lockdown are plotted in Fig 1 and Fig 2 respectively. In those plots, the trends were roughly linear and rational. It also depicts that there is little to no deviation of variance - conforming to a normal distribution of data. Although few of the data had shown a slight deviation, it could still be neglected due to the quantity being absolute meagre compared to the data which follows the trend.

**Fig. 1.**
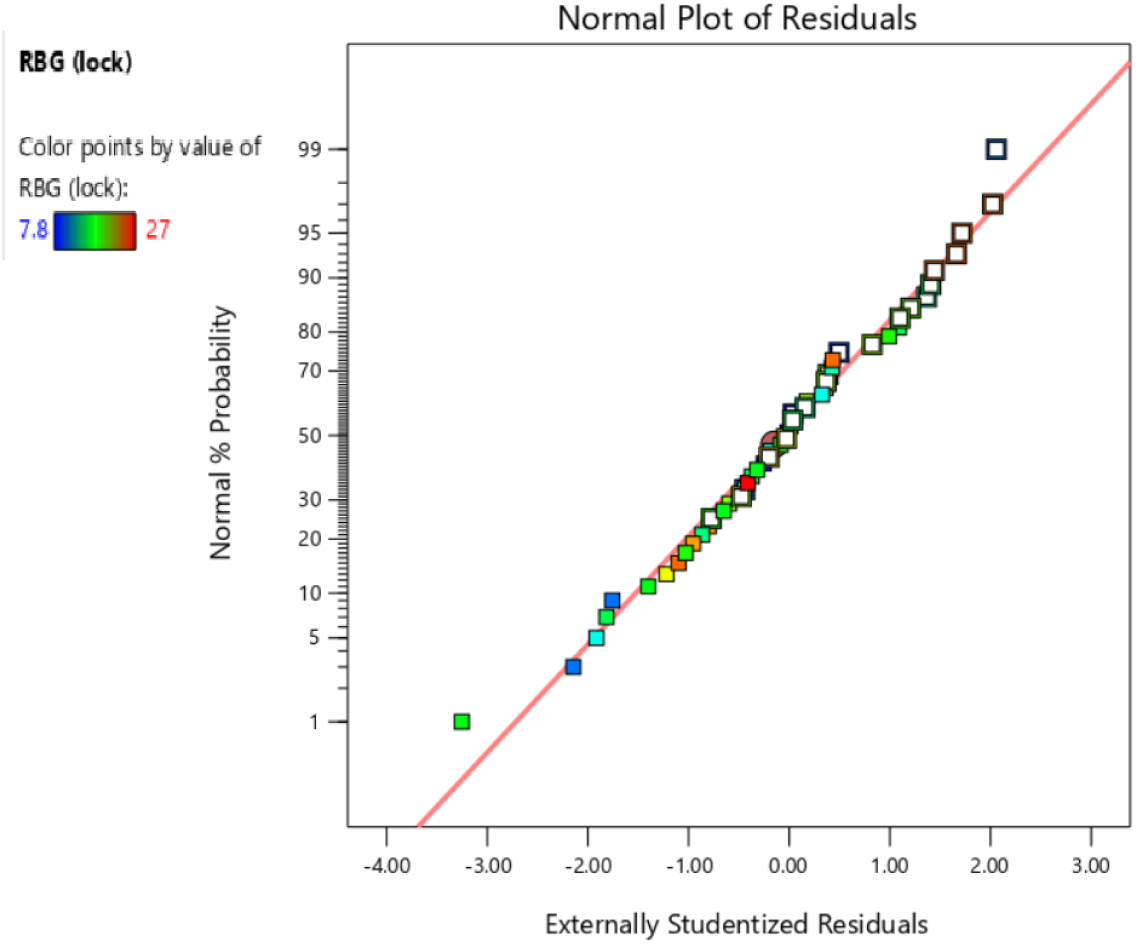
Normal probability plot for FBG during lockdown

**Fig. 2.**
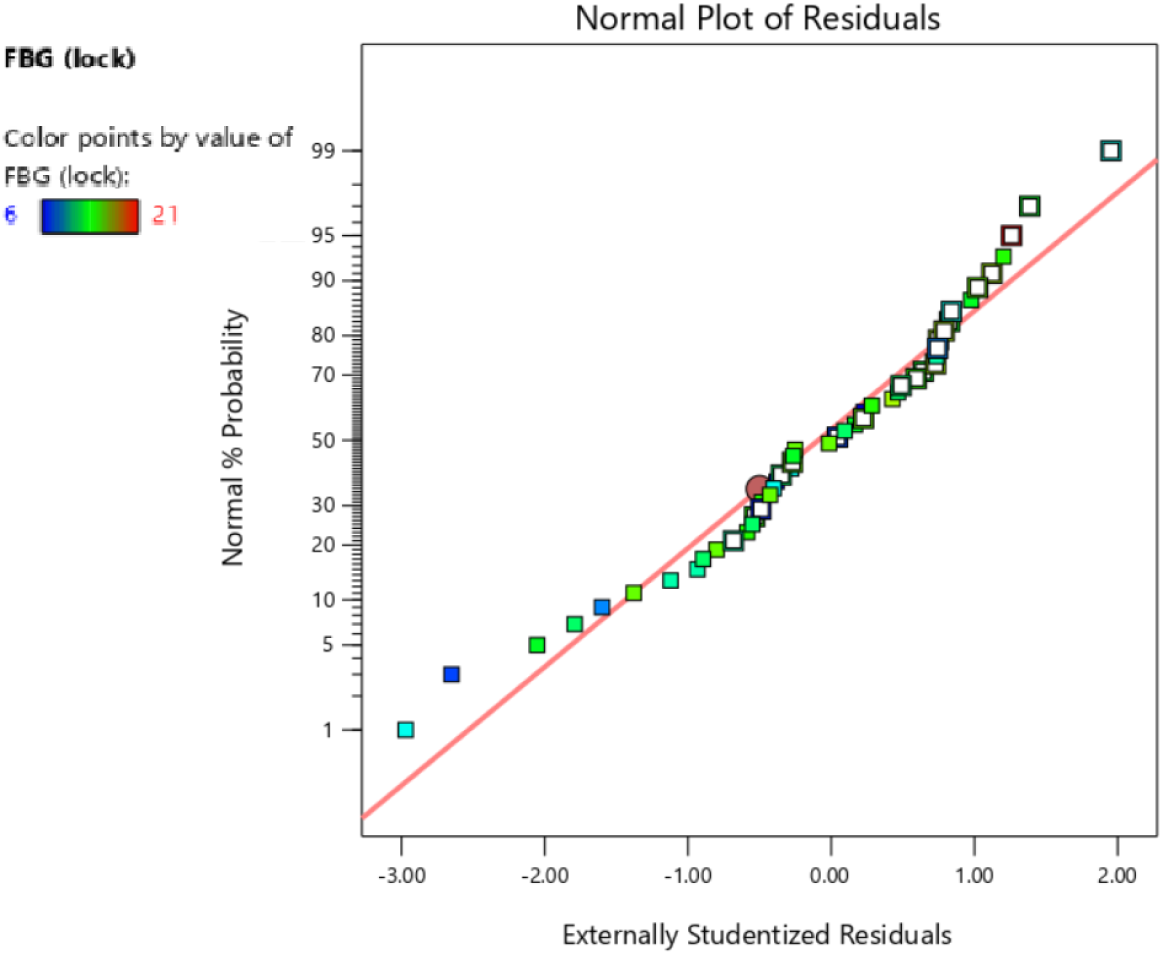
Normal probability plot for RBG during the lockdown

Predicted values were calculated - generated by Stat-Ease software - and compared with the empirical values collected from this study for RBG and FBG during lockdown. The predicted value generated by the model was through the prediction equation. The predicted value uses block and center point correction. In the equation, Y is the predicted value generated corresponding to the input data X. The ‘*β*’ symbol represents the gradient of the line.

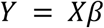

The comparison graph between predicted and actual values are shown in Fig 3 & 4. Both Fig 3 and 4 have shown a suitable correlation between predicted and experimental value. Adjacent to the straight line there are both values lying within the regression line and adjacent to it - marking a wonderful relationship. This agreement indicates that the regression equation has fitted the data appropriately with both FBG and RBG during the lockdown.

**Fig. 3.**
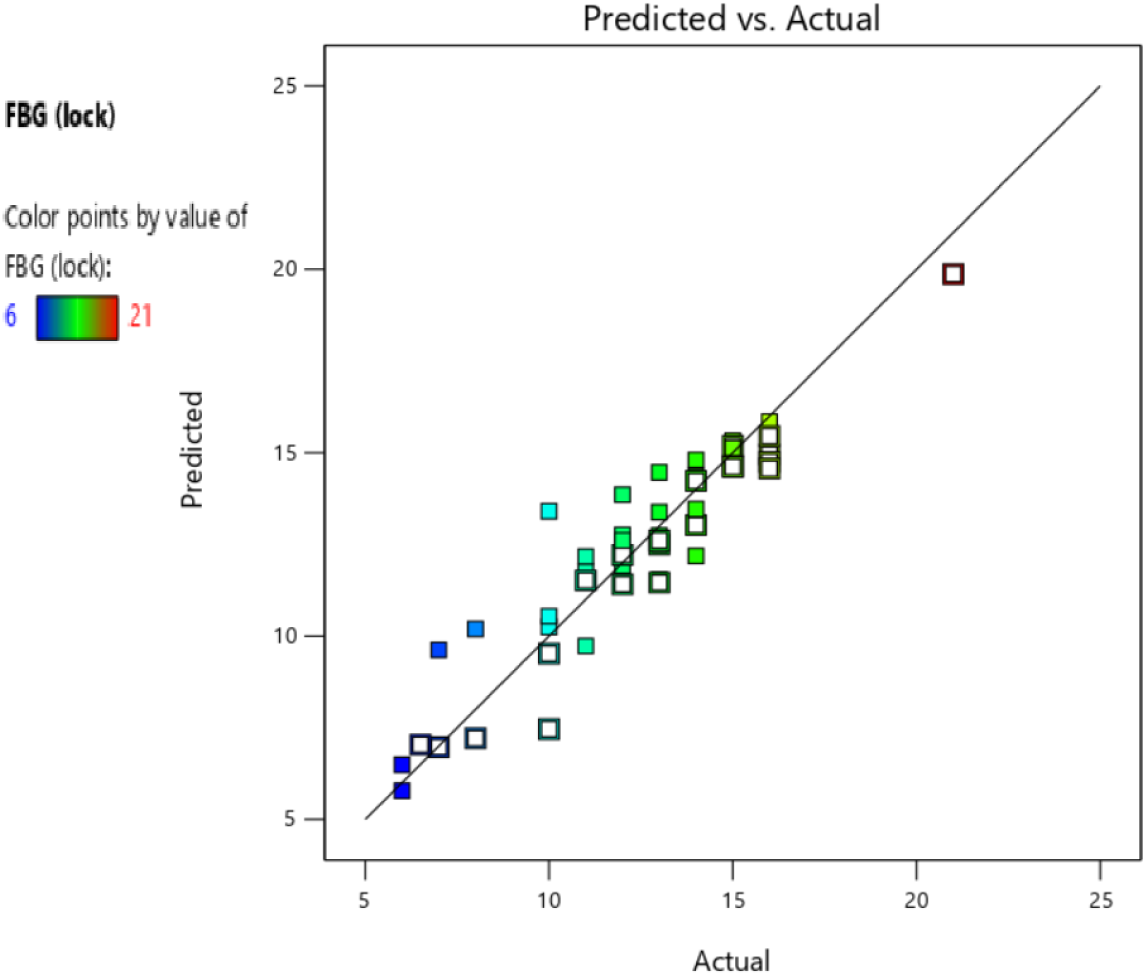
The linear correlation of actual and predicted response plot for FBG during the lockdown

**Fig. 4.**
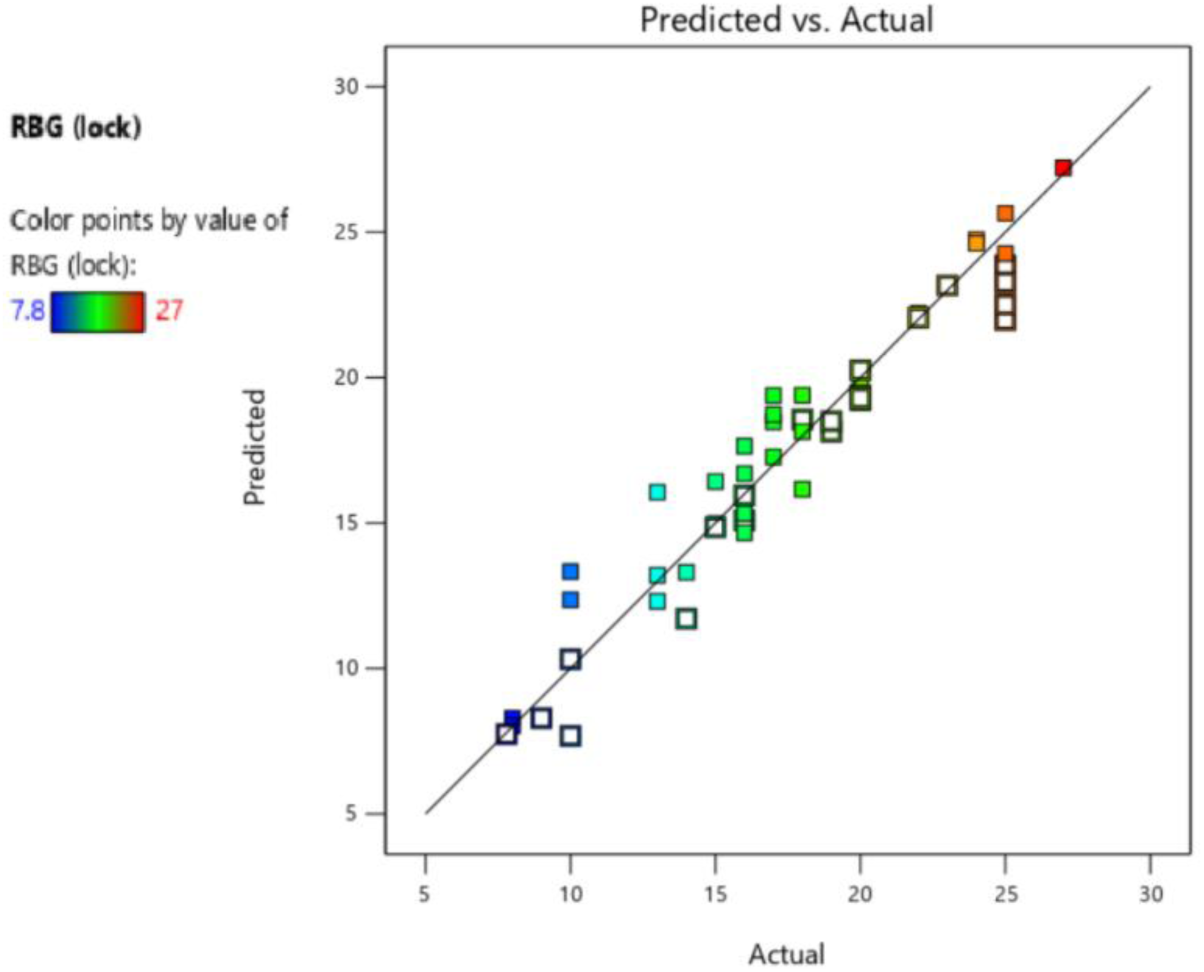
The linear correlation of actual and predicted response plot for RBG during lockdown.

In this research, the probability of the gaining error or any other possibility of error (such as outliers) for data acquisition was tested cautiously throughout. FBG and RBG quantities are shown in the “Residual vs. Predicted” plot in Fig 5 and 6 respectively. The degree of standard deviation was calculated from this plot. This plot indicates the intensity of deviation between actual and predicted value. The range of maximum standard residuals is ±4.00 required. Any value beyond this standard residual value was discarded from the experimental response. From this study, it is observed that almost all are lower than the outlier. These two Figs are justifying for this research that this has fitted the model without any influenced error. The random scatter of data in the plot depicts that the empirical data had a constant variance.

**Fig. 5.**
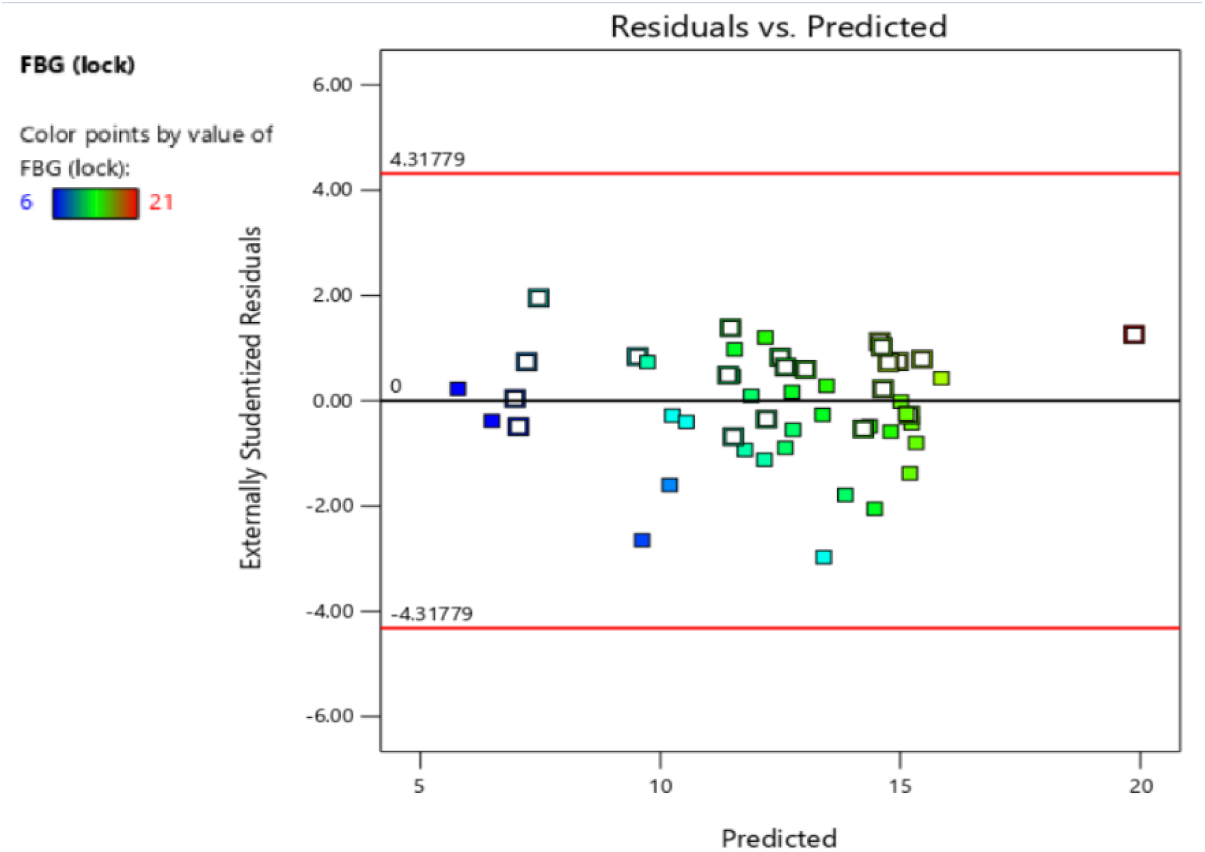
The residual and predicted response for FBG during the lockdown.

**Fig. 6.**
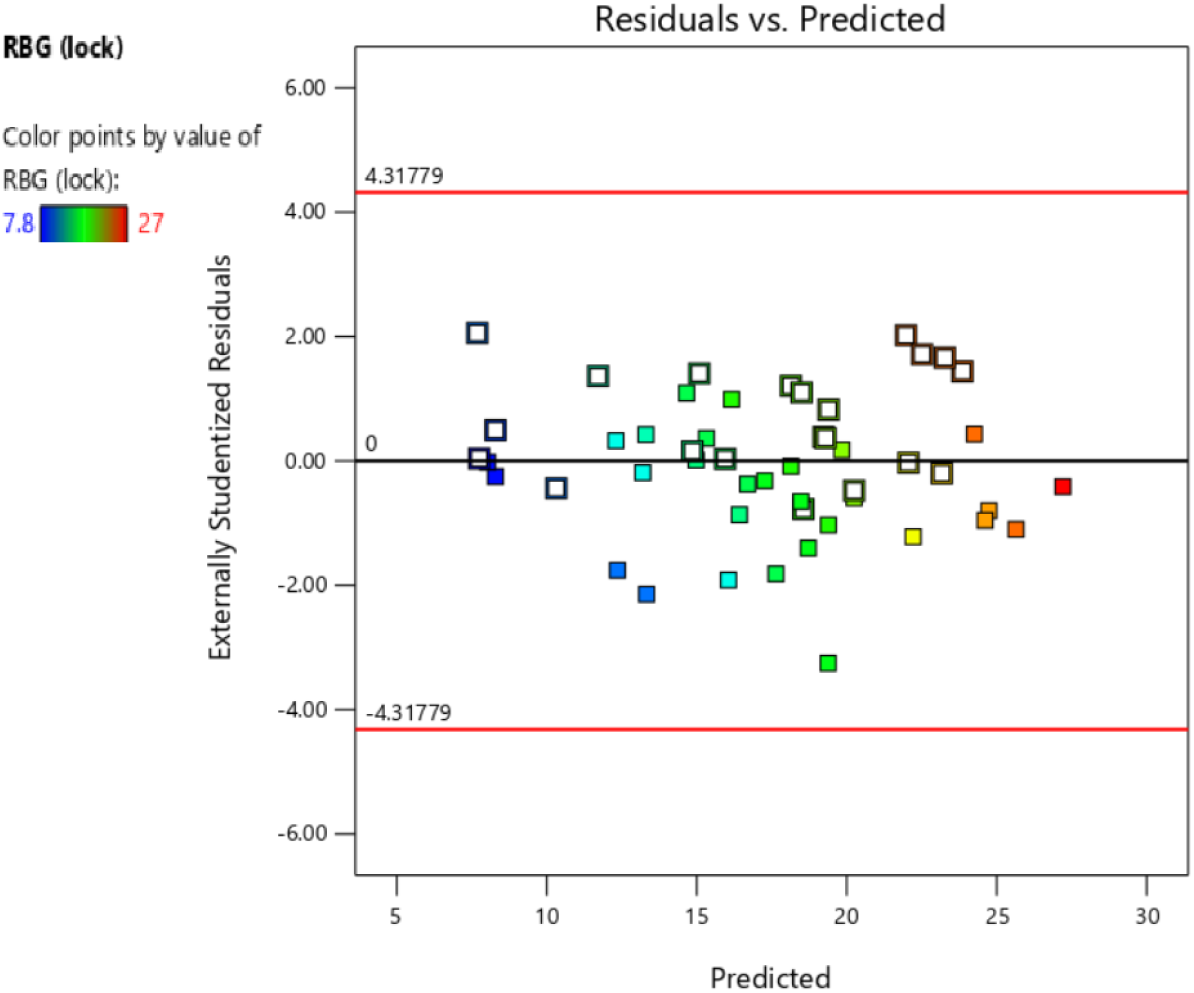
The residual and predicted response for RBG during the lockdown.

Stat-ease software offered different power transformations that could be applied to improve the model and yield the most accurate statistical analysis. As the response vectors were all greater than zero, power transformation could be applied. The software had generated the best power transformation that could be applied before ANOVA was conducted. The selection process of the optimum power transformation was determined using the ‘lambda’ value generated - the minimum point of the curve generated by the natural log of the sum of squares of the residuals. In this statistical analysis (for both FBG and RBG analysis), the 95% confidence interval lies around 1. Thus, there wasn’t any need to apply power transformation. The Box-Cox plot generated by the software had shown this result and suggested that no transformation is required for the statistical analysis. The Box-Cox plot for FBG and RBG is shown in Fig 7 and 8 respectively.

**Fig. 7.**
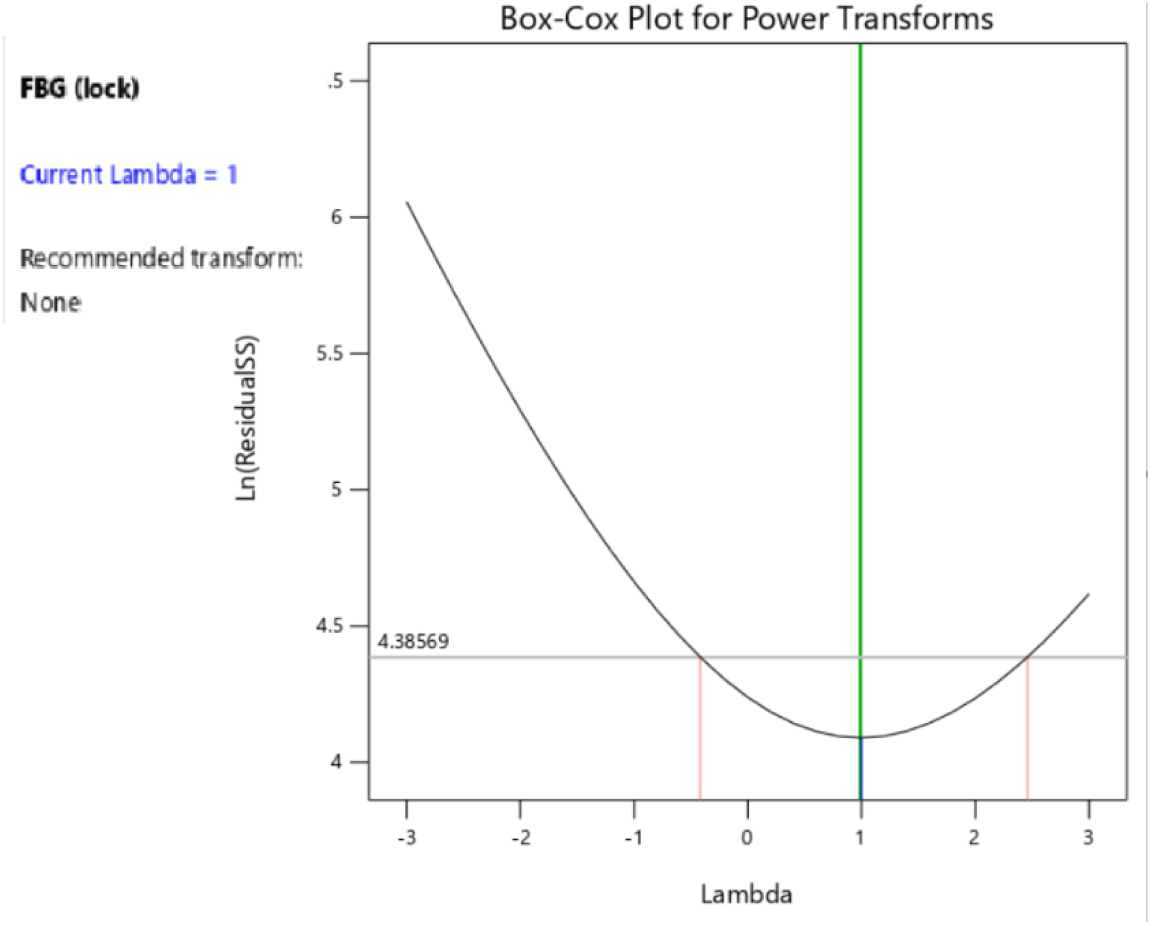
The Box-Cox plot for FBG during the lockdown.

**Fig. 8.**
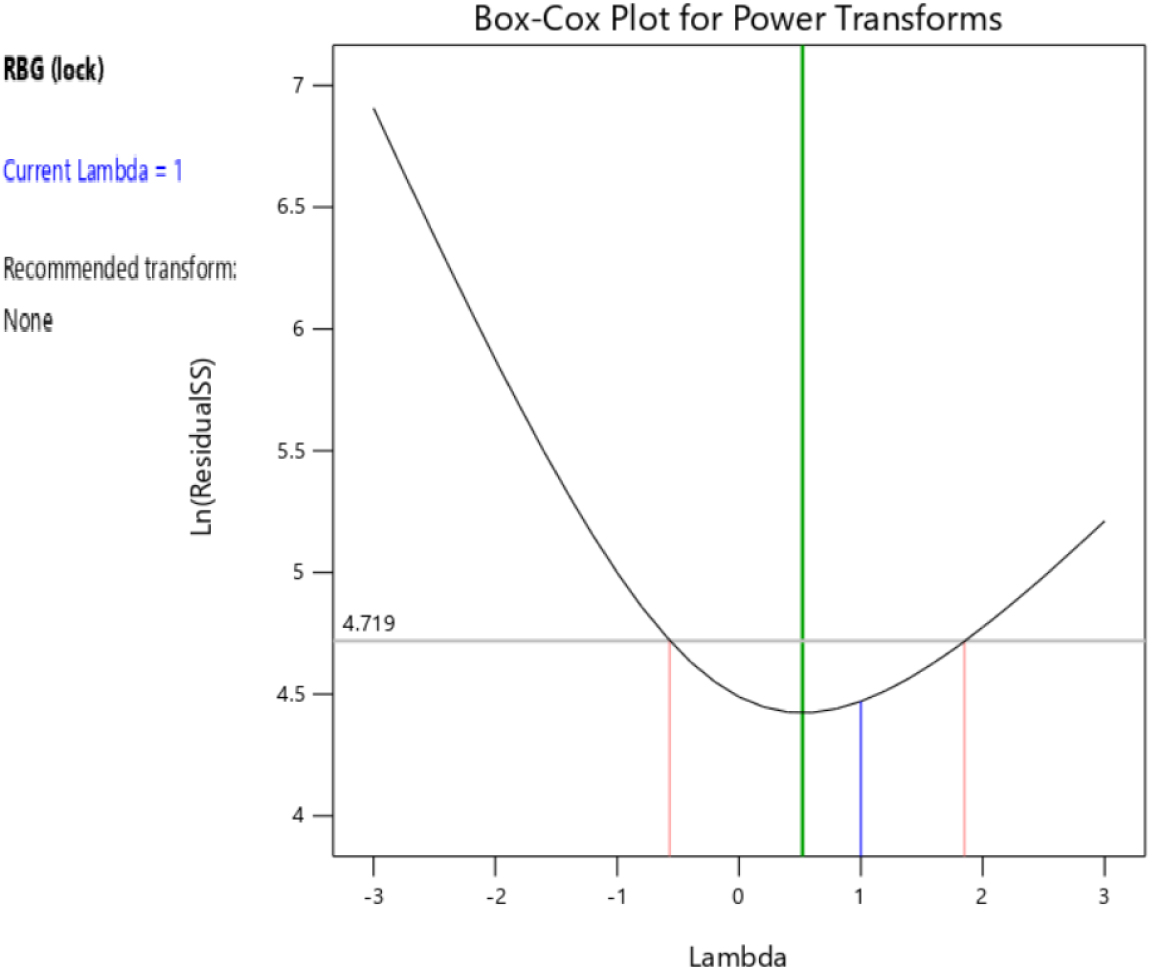
The Box-Cox plot for RBG during the lockdown.

## 3. Results

### Model Graph and Evaluation

RSM renders numerous benefits such as the interaction effect of independent parameters and detecting the spike in BGL in the effects of the binary combination of linking two independent parameters. From the graph, it is easier to understand the relation between different factors. For graphical explanation, three-dimensional plots are very useful. Equation 6 and 7 were used to plot the 3-D response surfaces. This is showing that the FBG and RBG were affected during the lockdown and after the lockdown with various parameters.

The perturbation diagram for FBG and RBG after lockdown is shown in Fig 9 & 10 respectively with respect to the seven input process factors. The process variables having influence around a specific point is illustrated in the design range shown in this perturbation plot. FBG and RBG were plotted with respect to one variable from the overall process. Design Expert has automatically set the reference point at the midpoint, which is coded 0, of all the factors. At the center point, additional process variables remain constant. When the response is sensitive then a steep line is observed, and a flat line is representative of insensitive factors. In the two generated perturbation plots each feature tested for the response vector is represented by specific symbols. For both Figs 9 and 10, the coded perturbation graph, the left image is the result for the female patients and the right image is the result for the male patients. From Fig 9, it can be interpreted that most of the factors were relatively flat for the female patients in their rise of the BGL, but it had shown a steep slope on the SBP and RBG as factors. On the other hand, the male patients showed various steep slopes corresponding to their factors - indicating their rise in blood glucose being sensitive to the factors. In Fig 10, the perturbation graph of both male and female had a steep slope for most of the factors - this indicates that the rise in the BGL among the patients was affected by these factors. From both Figs, it could be depicted that the male patients had slightly higher FBG and RBG values during the lockdown.

**Fig. 9.**
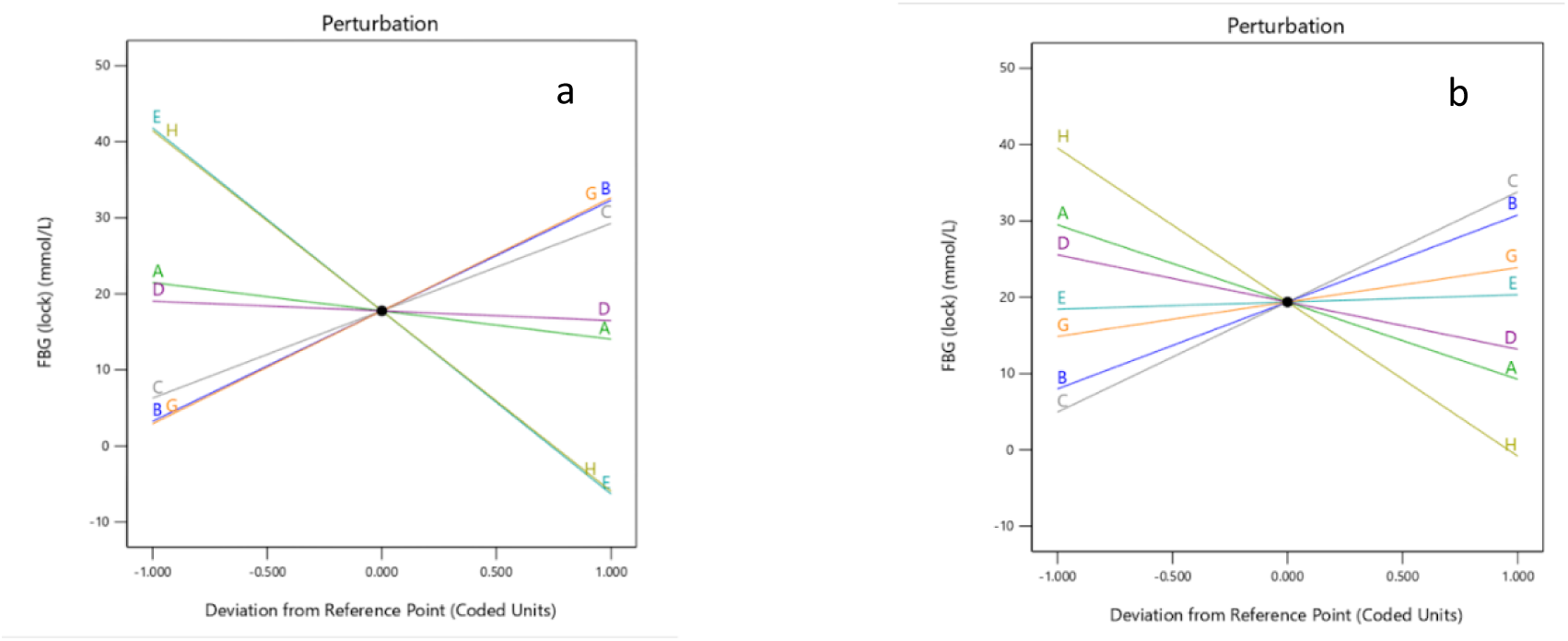
Perturbation graph of FBG for male and female (mean).

**Fig. 10.**
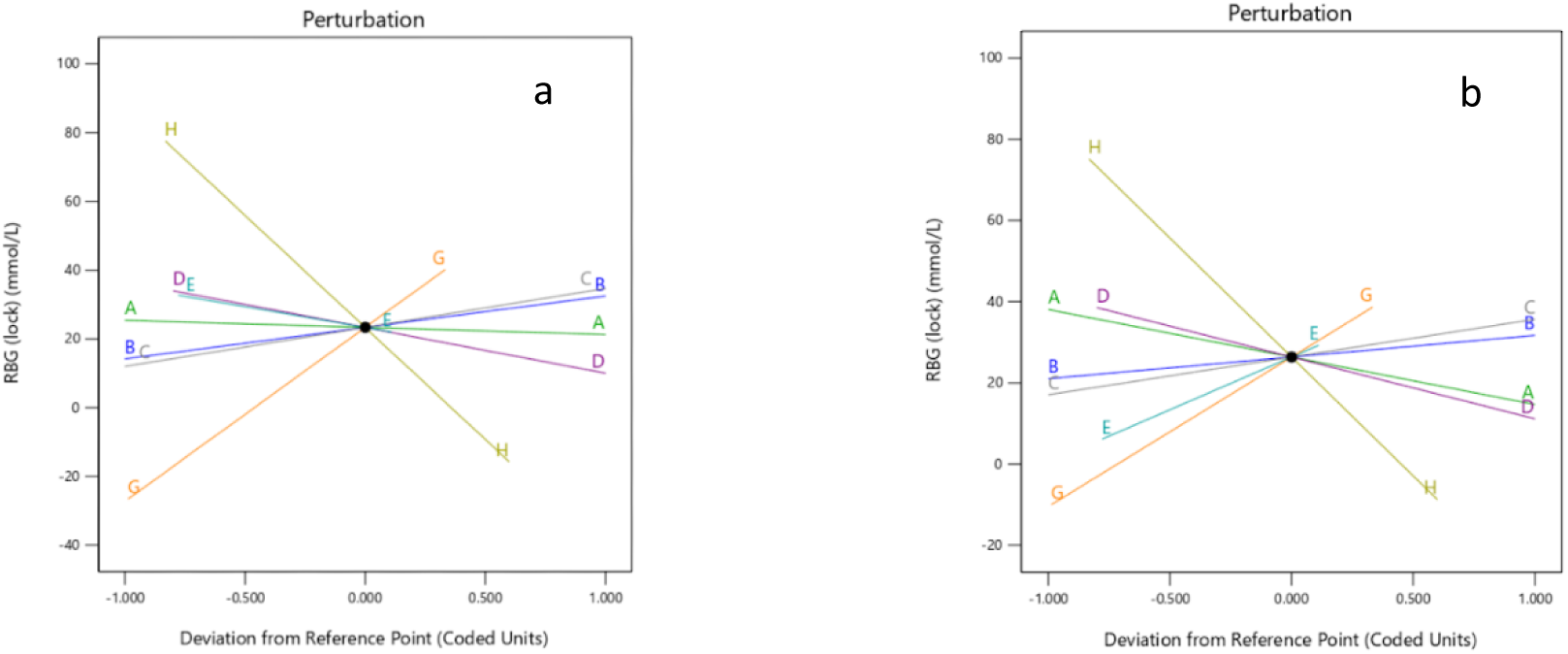
Perturbation graph of RBG for female and male (mean)

The 3D Response surface can explain the interactive characteristics of features. To mark the interaction between the RBG and FBG after lockdown, 3D graphs are used. The surface highest point is indicating the highest level. In the Figs, various effects are indicated by color lines on the FBG and RBG increasing rate - the red portion indicates the highest results. Many 3D graphs were generated by the Design-Expert software, but in this analysis, only the most prospective relations are demonstrated. In Fig 11, the 3D graph shows the interaction between the BMI of the patient and the DD among patients - for both FBG and RBG analysis. BMI was illustrated on the X1 axis and DD was illustrated on the X2 axis. The graph showed an average over result between male and female patients. On the right side of the graph, the FBG (lock) graph is shown and RBG (lock) on the left side. The graph was set upon the mean values generated in Table 1 yet it shows an overall response surface and the interaction among the parameters. In the graph, it is seen that the patients with high BMI and DD were prone to have a high-level during lockdown - for both RBG and FBG. However, if a patient has a high BMI and DD then their FBG is more prone to rise than RBG: due to the steeper peak in the FBG curve.

**Fig. 11:**
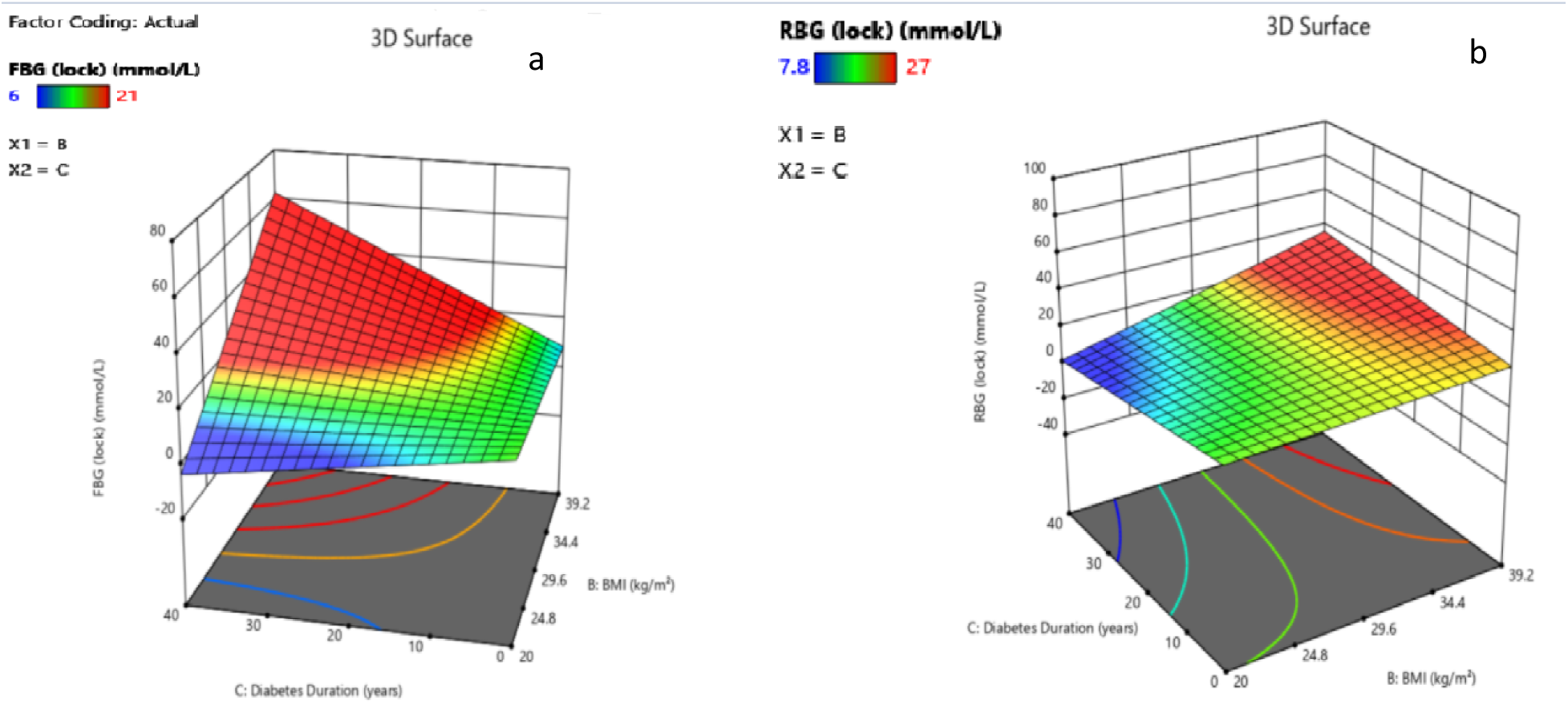
3D surface graph of BMI and DD.

Another special type of graph plot was generated, and it is known as the “Contour Plot”. It is a two-dimensional graph used to show the interaction between two different features with the response vector. In Fig. 12, the contour plot analyzing the effect of BP on BGL is analyzed for both FBG and RBG. In the plot, the X1 axis contains SBP and the X2 axis contains DBP. Once again, the plot was generated as an average over between male and female, and it was initially set with the mean value from Table 1. On the right-hand side of the plot, the FBG plot is illustrated, and on the right-hand side of the plot, the RBG plot is illustrated. From the plots, an interesting perspective is presented, the DM patients who had high SBP were rather prone to high FBG during the lockdown. On the contrary, the patients who had high DBP values tend to have high RBG during the lockdown. This clearly shows that the blood pressure level of patients is sensitive to the rise in BGL.

**Fig. 12.**
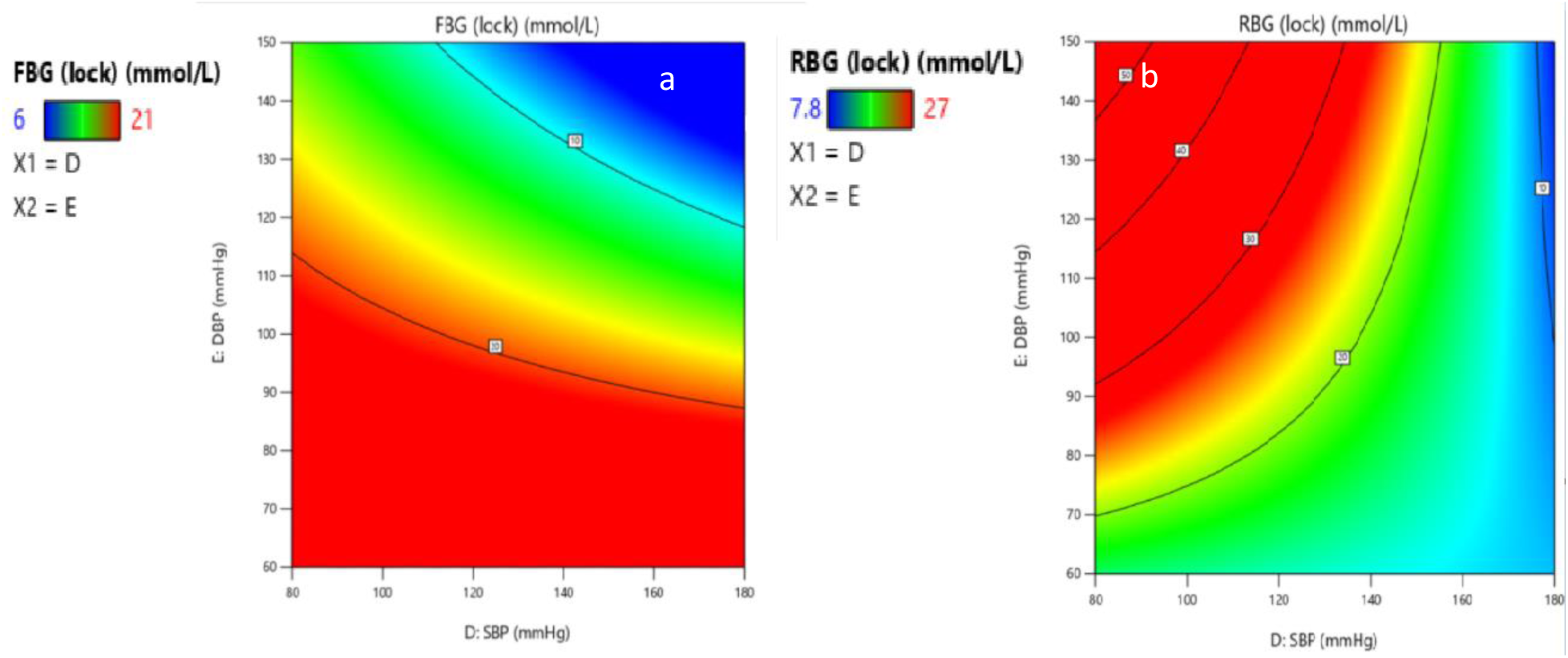
Effect of BP on the rise of BGL.

Thirdly, from the 3D graph of figures 13 the relationship between the age of the patient and the RBG before lockdown in response to the RBG during lockdown is established. In the X1 axis of the graph, age is illustrated and on the X2 axis, the RBG before lockdown is illustrated. Again, the graph was generated based on the mean value of each factor from Table 1. In addition, the graph was generated in average over the result between male and female patients. The graph demonstrates the elderly patients who had high RBG before lockdown were prone to rise in RBG during the lockdown. As well as, the younger patients were less prone to the rise in RBG than the elderly patients during the lockdown. Similarly, if a patient had low RBG before the lockdown they were less likely to have a spike in the RBG during the lockdown.

**Fig. 13.**
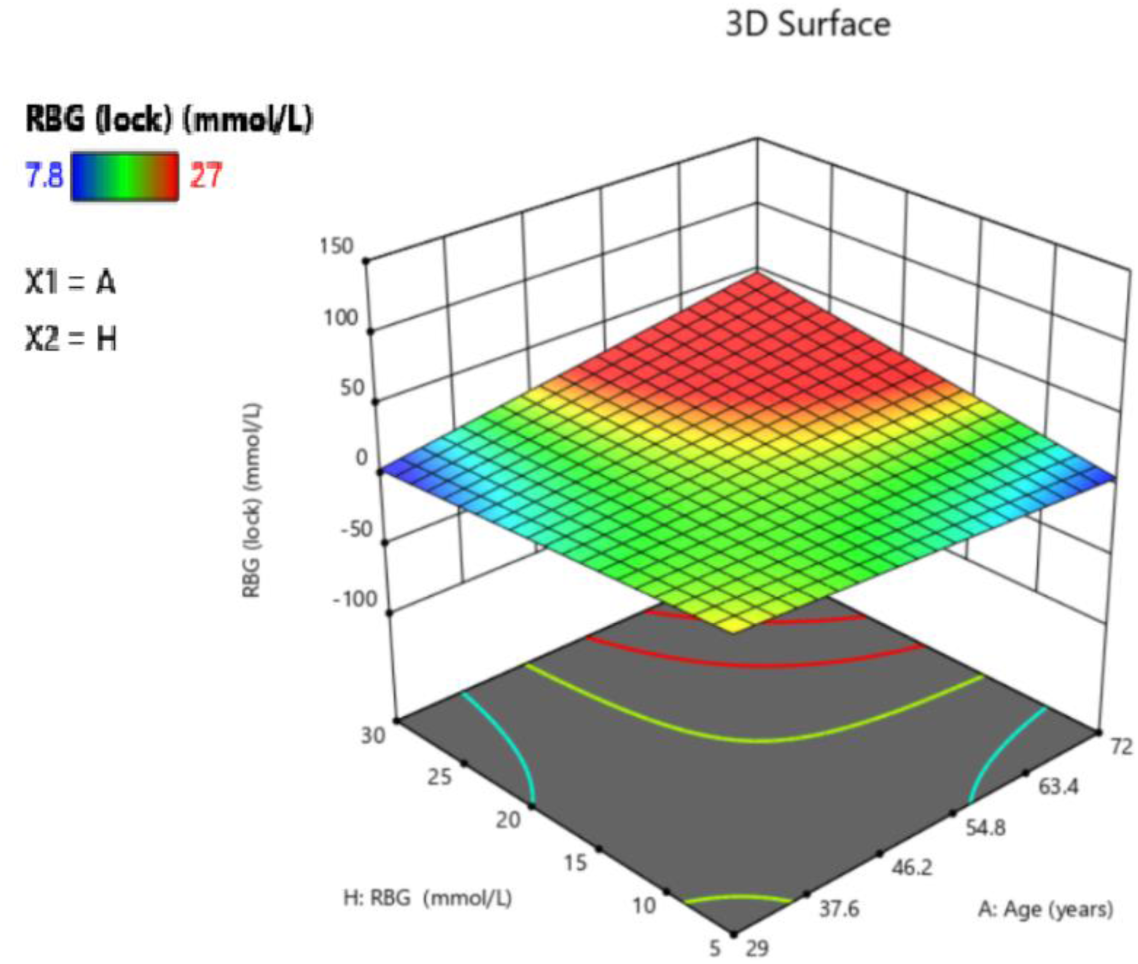
Effect of Age and RBG before lockdown on the rise of RBG during the lockdown.

## 4. Discussion

### The effect on the economy due to SARS-2

As a result of high BGLs of DM patients during the lockdown, they are subjected to many complications leading them to frequent visits to hospitals. This adverse health effect will have a negative impact on the economy [21-25]. A study conducted by Afsana Afroz et al. pointed out that DM patients of Bangladesh who have three or more complications spend approximately three times more than the healthy DM patients [13]. In other studies, it had shown that the cost of maintaining DM increased with the number of comorbidities and complications the patients faced. These findings were supported in both developed [26-28] and in developing countries [29-30].

Firstly, DM imposes a large financial burden on patients and their families. People in developing countries have to pay a large share of their expenditure on medical services. Uncontrolled sugar levels will affect individuals, families, providers of medical care and as well as the ministry that pays for care. Then the national economic growth will be hindered. Below an estimation of expenditure due to the hyperglycemia during the lockdown is given, depicting the rise in expenditure for patients. Below Table-5 is given as an approximation of expenditure of DM patients after lockdown. Table 5: Approximation overall expenditure of DM patient’s post-lockdown.

**Table 5:**
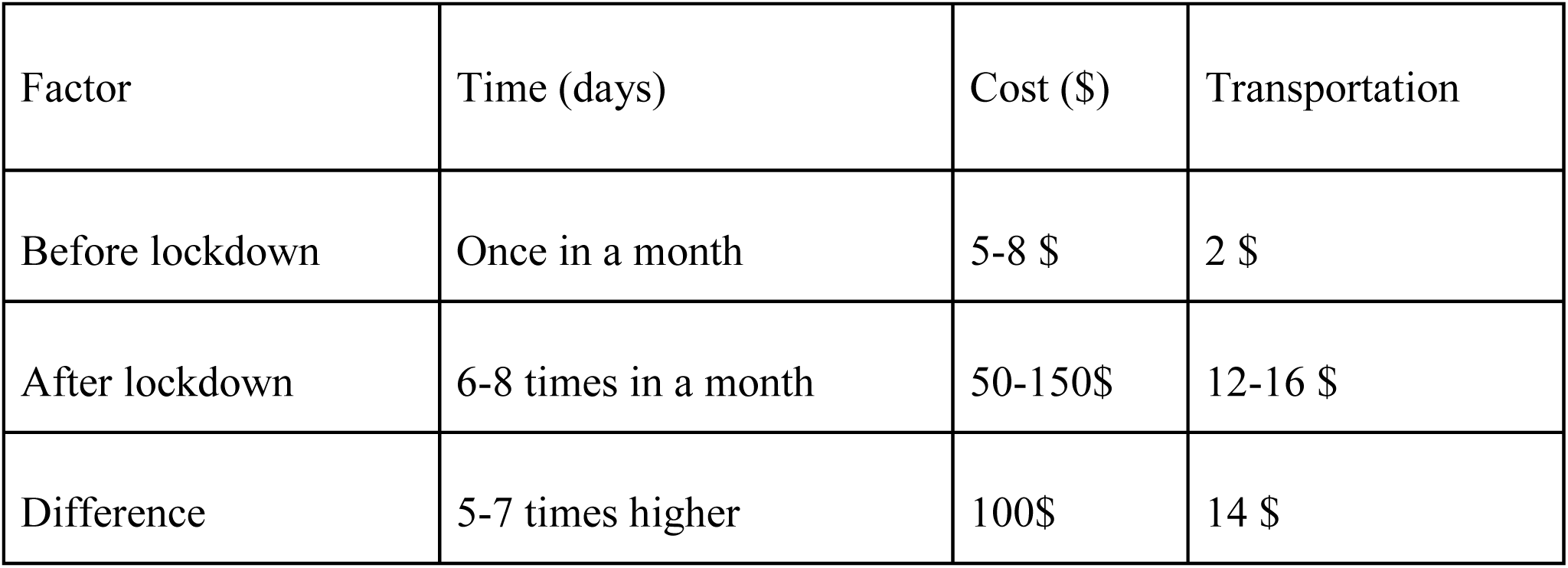
Approximation overall expenditure of DM patients post-lockdown.

| Factor | Time (days) | Cost (\$) | Transportation |
| --- | --- | --- | --- |
| Before lockdown | Once in a month | 5-8 \$ | 2 \$ |
| After lockdown | 6-8 times in a month | 50-150\$ | 12-16 \$ |
| Difference | 5-7 times higher | 100\$ | 14 \$ |

## Conclusion

In this statistical analysis, the author tried to analyze the factors associated with the upsurge in BGL during lockdown by applying the sequential sum-of-squares model (2FI). The model applied is significant as it had shown *p*-value of 0.0441 and 0.0015 for the two response vectors - this justifies that the null hypothesis could be avoided. In addition, the *F*-value of the applied model in FBG(lock) and RBG (lock) response vectors were 2.43 and 5.1 respectively. ANOVA method applied to select the model, power transformations and diagnostics tools. The detection of outliers among the dataset was conducted using diagnostics tools such as predicted vs residual plot. The *R-*squared values used for FBG (lock) and RBG (lock) were 0.87 and 0.94 respectively, justifying that the regression was fit to the dataset. Next, the RSM applied to yield out the interactions among parameters. Subsequently, it is found that the male patients were subjected to higher levels of BGL during lockdown than the female patients. As well as, the patients with high BMI level and diabetes duration have high BGL during the lockdown. On the other hand, if an elderly patient had a high level of RBG pre-lockdown, then he/she was more likely to have a high level of RBG during the lockdown. Also, if a patient had high SBP, then it was likely that he/she would have a high FBG level during the lockdown. If a patient had a high DBP level, then he/she was subjected to a high level of RBG during the lockdown. Although the sample set didn’t comprise an extensive number of patients, it was still unique as it had 8 total factors and 2 response vectors, and the relationship among the parameters was analyzed with the RSM. This survey was taken recently after the pandemic lockdown in 2021.

## Data Availability

The data will be available when requested.

## Availability of data and material

The data will be available when requested.

## Notations

*SSQ*: sum of squares
*SSQ*_*model*_: sum of squares of model
*SSQ*_*residual*_: sum of squares of residual
*p*: number of model parameters
n: number of experiments
*ANOVA*: analysis of variance
*Mn*_*SRG*_: mean of square regression
*Mn*_*SRD*_: mean of square residual
*MnSer*: Mean Square Error
*SRG*: sum of squares
*SRD*: sum of residual
*Df /DgF*_*mod*_: degree of freedom of model
*DgF*_*residual*_: degree of freedom of residual
*R*^*2*^: determination coefficient
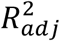: coefficient of determination
*CV*: coefficients of variation
*F-value*: model significance
*SD*: Standard Deviation

Click here to access/download

**[Supplementary Material](https://www.editorialmanager.com/heliyon/download.aspx?id=754187&guid=9f6f5e98-f0e6-424a-b282-7b26d13573b3&scheme=1)**

Ethical Approval Statement.pdf

